# Prevalence and associated occupational factors for low back pain among the bank employees in Dhaka City

**DOI:** 10.1101/19012328

**Authors:** Mohammad Ali, Gias U. Ahsan, Ahmed Hossain

**Affiliations:** Department of Physiotherapy, Uttara Adhunik Medical College Hospital, Uttara, Dhaka-1230, Bangladesh; Centre for Higher Studies and Research, Bangladesh University of Professionals, Dhaka-1216, Bangladesh; Department of Public Health, North South University, Bashundhara, Dhaka-1229, Bangladesh; Health Management BD Foundation, Dhaka-1230, Bangladesh

**Keywords:** Occupational Health, Low Back Pain, Bank employees, random forest, Bangladesh

## Abstract

**Objective:** Low Back Pain (LBP) is one of the common health problems among full-time office employees that causes employees absenteeism from work. The purpose of the study is to identify the association between occupational factors and LBP among full-time bank employees in Dhaka City.

**Materials and Methods:** We conducted a cross-sectional study with 593 full-time bank employees. The one-month complaints of LBP were administered by a musculoskeletal subscale of subjective health complaints by Eriksen et al. A logistic model was performed to identify variables associated with LBP, and a random forest technique was performed to identify the top 5 important variables.

**Results:** The one-month prevalence for LBP was found 36.6% among the bank employees and the prevalence was high (46.6%) for the 41 to 59-year-old age-group. The multiple logistic regression analysis indicates that age (41-59 years) (OR:2.11, CI=1.21-3.74), obesity (OR:2.06, CI=1.01-4.21) and long working hours (>9 hours) (OR:1.42, CI=1.01-2.0) are positively associated with LBP. Age and length of employment have a positive correlation of 0.87. The random forest technique identifies the top 5 important variables are, age, length of employment, long office hours, presence of chronic illness, and physical activity.

**Conclusion:** LBP is highly prevalent in full-time bank employees. The occupational factors like length of employment (>10 years) and long working hours (>9 hours) play a significant role in developing LBP among the bank employees. Moreover, the factors like age, chronic illness, obesity and physical activity should be taken into account in the prevention of LBP in bank employees.

## Background

The global burden of diseases, injuries, and risk factors study in 2016 suggested that among 328 morbidities, low back pain (LBP) became one of the most significant health concerns for any population group [1]. Another study showed that the lifetime prevalence of LBP reached up to 84% [2]. LBP can induce a lack of enthusiasm, mental unrest, and physical discomfort or burden on its bearer [3]. Consequently, LBP became a significant cause of taking sick leave and early retirement among the working population [4].

Recently non-manual occupations made essential changes in our working life [5]. Office workers spend a substantial amount of time sitting at a desk. Recent studies have shown that the prevalence of LBP among office workers varies from 34% to 51% [6-8]. The prevalence of LBP in low-income countries were found higher compared to high-income countries [9]. It is, therefore, essential to explore the prevalence rate of LBP in Bangladeshi full-time office workers. A study revealed that long-time sedentary work, high workload, and inappropriate sitting arrangements are the contemporary causes of LBP [10]. Many studies conducted with office workers found a relationship between sitting and LBP [6,11-15]. Another study suggested that the associated factors of LBP for office workers were: long office hours, working in the same posture, and continuing the same job for many years [3]. Furthermore, few studies revealed that prolonged sitting was associated with metabolic disorders, sleep disturbance, hypertension, and high body mass index (BMI) [16-17]. These factors are also positively associated with increased LBP [18]. Studies on work-related LBP in Bangladesh found high prevalence among different work settings. For example, female garments workers (34.6%) [19] and professional female nurses (31.8%) reported high prevalence of chronic LBP [20]. Another study conducted by Hossain et al. on work-related musculoskeletal disorders found a 24.7% prevalence of LBP among garments workers of both sexes [21]. However, there is a research gap in evaluating the effect of work-related factors and low back pain in the growing number of full-time office employees in Bangladesh. Thus, this study aims to determine the prevalence of LBP and find associated occupational factors among office employees in Dhaka city.

## Methods

We conducted an analytical cross-sectional study in Dhaka City between December 2018 and May 2019. Dhaka is the capital city and economic hub of Bangladesh. We selected full-time bank employees who maintained a regular office hour for at least one year at a bank. There are 50 banks in Dhaka City, and we conveniently selected 32 banks to collect data from their full-time employees. We included the employees who were working in the bank for the last year. Figure 1 represents the flow chart of the data collection. We distributed 923 paper-based questionnaires to the employees during office hours, and we collected 652 questionnaires during the study period. After scrutiny, we found 628 completed questionnaires. We excluded nourishing mothers, pregnant women, and those who were bearing chronic inflammatory pain (e, g., rheumatoid arthritis, ankylosing spondylitis). We also excluded employees who were working while standing. After taking into consideration of the inclusion and exclusion criteria, we took 593 participants in our analysis.

**Figure 1:**
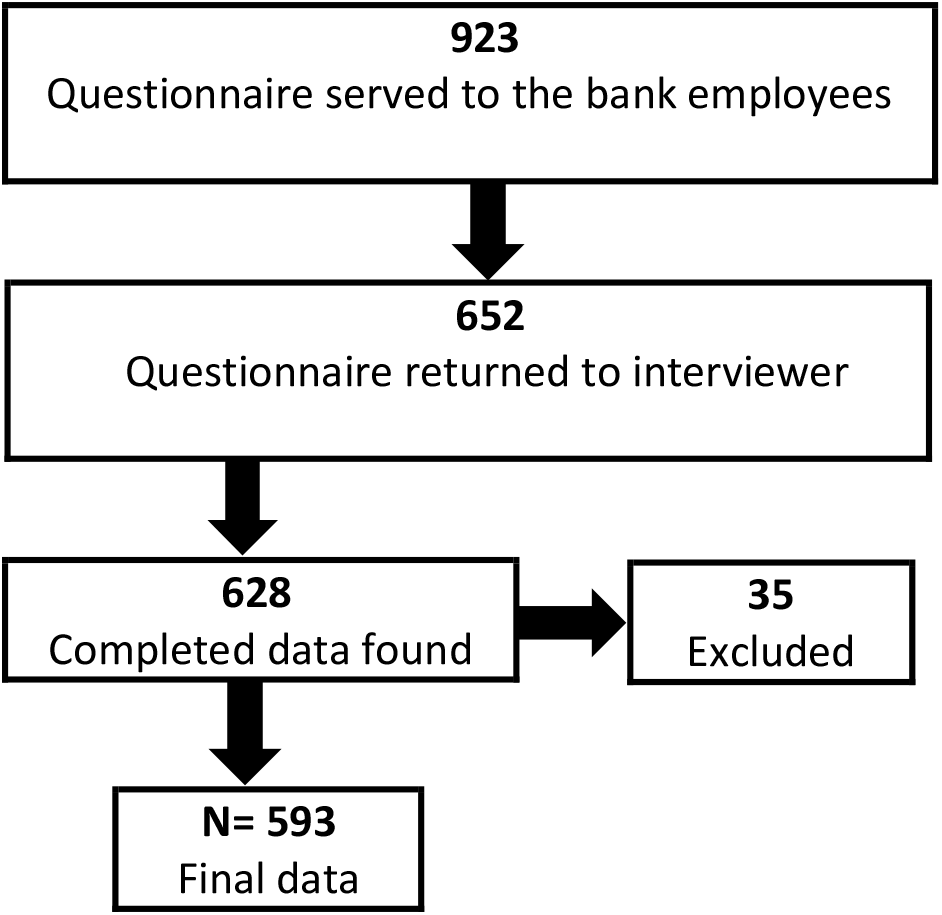
Flow chart of data collection.

### Instruments

The questions on LBP were based on the musculoskeletal subscale of subjective health complaints produced by Eriksen et al. that measure LBP complaints experienced in the last month [22]. Employees were asked to rate the occurrence of pain or discomfort in the lower back with four answering categories (“no complaint,” “only once/a little,” “of short duration/ some,” “frequently/ serious”). Employees who answered, “no complaint,” “only once/a little,” “of short duration/ some” on LBP were classified as having no low back pain. Those who answered “frequently/serious” were classified as having complaints of LBP.

Data on socio-demographic factors-age, gender, BMI (calculated based on weight and height), and marital status were collected using a semi-structured questionnaire. Behavioral factors like sleep arrangements, smoking habits, and physical activities of the respondents were collected. Physical activities were calculated based on the METs scale [23]. We also collected occupational factors like length of employment and average daily working hours. The crowding was calculated by dividing number of family members in the house by the number of bedrooms in the house. Data regarding common chronic illness (Diabetes and Hypertension) from the employees were also collected.

## Result

### Univariate Analysis

Among the 593 respondents, there were 342 (57.7%) males and 251 (42.3%) females. The descriptive statistic of the factors such as age, gender, BMI, marital status, crowding, and sleeping habits, are described in Table-1. More than half of the participants (59.02%) were between 31 and 40 years of age, and about half of the employees were overweight or obese.

The one-month prevalence of the complaints about LBP is found to be 36.6% among the bank employees. There was no difference in prevalence of LBP between the gender, but had influence of age and body mass index. The results of the p-value from the chi-square statistic indicates that age has a significant association with LBP. In this study, the majority of the participants were married (83.8%), and more complaints of LBP from married employees were found compared to unmarried participants. It is because more aged employees were married. Thus, to avoid the collinearity in the multiple logistic regression model, we excluded marital status in the model. The results indicate a high prevalence (48.4%) of LBP in the 41 to 59-year-old age group, indicating that it is a common condition among older adults.

The behavioral and occupational factors of the participants, such as smoking habits, chronic illness, physical activity, length of the employment in a bank (years), and average working hours per day are described in Table 2. More than half of the participants (56.2%) maintained regular office hours (8-9 hours) per day. However, those who continued extended office hours (spent more than 9 hours in the office per day) complained more about LBP than the employees who maintained regular office hours (41.5% and 32.7%, respectively).

A scatter plot is shown in Figure 2 to understand the relationship between age, length of the employment, and complaints about LBP. The complaints about LBP also increased among full-time employees, as the length of the employment in the bank increased. The correlation between age and length of the employment in a bank is found 0.87, which indicates a high relationship. Therefore, to avoid multicollinearity, we included age in the multiple logistic regression analysis and excluded length of employment. Moreover, 46.6% of employees who had any chronic condition reported suffering from LBP (p=0.009).

**Figure 2:**
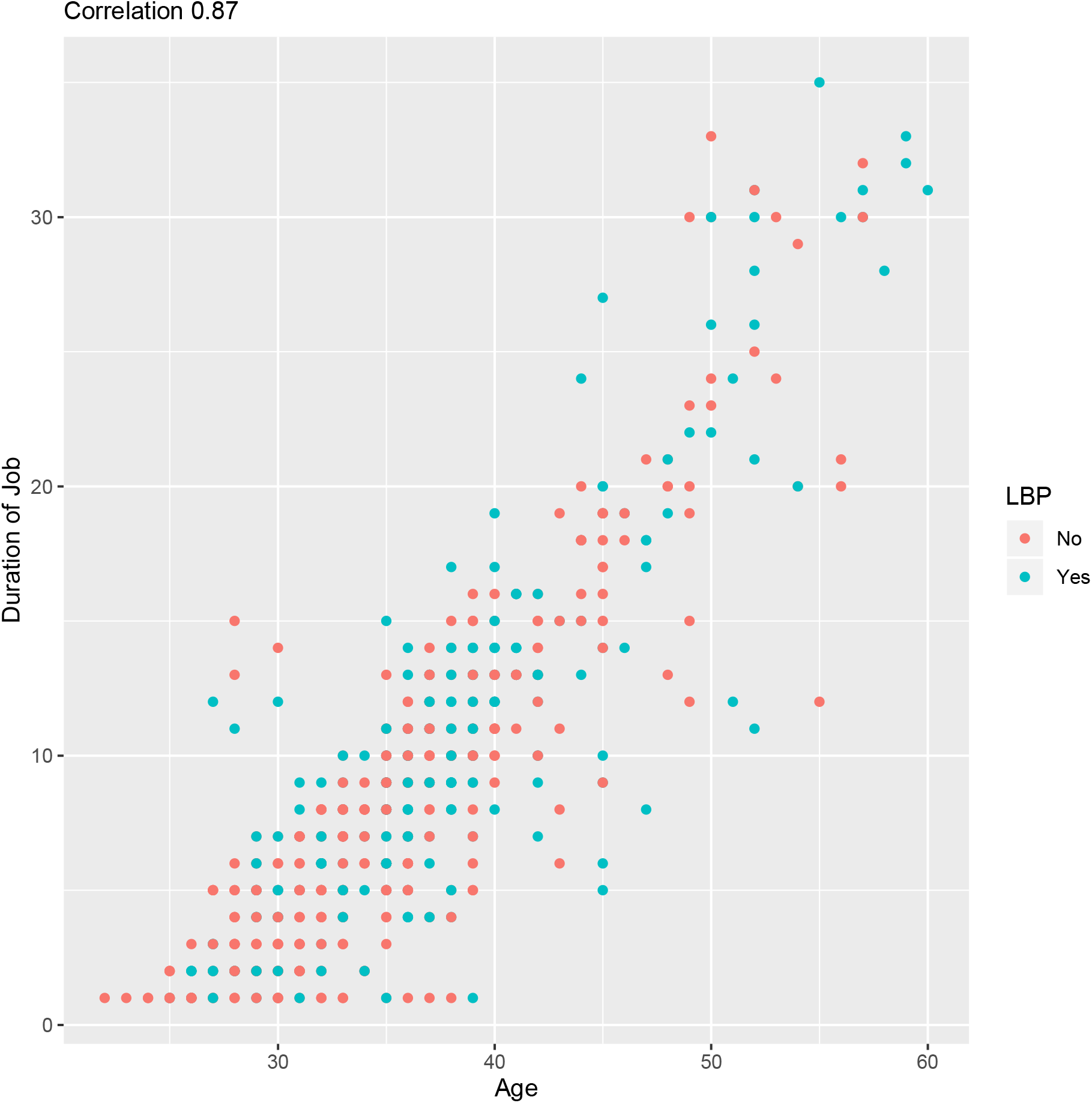
Correlation.

**Figure 2: Scatter plot to understand the relationship between age, duration of the employment in banks and complaints of LBP. It appears that most red dots are in the bottom which indicates new and younger employees are complaining less about LBP**.

### Logistic Regression Model

We fit a logistic regression model with the complaints of LBP after adjusting all the individual associated factors from Table 1 and Table 2. It appears from Table 3 that there are three factors associated with the complaints of LBP at 5% significance level. The three factors are: long working hours per day (OR=1.42), older age group (OR=2.11), and BMI category-obese (OR=2.06) were found to have an association with LBP. The results indicate that the bank employees who work for an extended period are 1.42 times more likely to have the LBP than those who work regular hours. Moreover, the age group of 41-59 years is 2.11 times more likely to have LBP than those in age group of less than 30 years. Obesity (OR= 2.06) is found to be significant in the development of LBP, which indicates an obese employee is 2.06 times more likely to have LBP than an employee from a healthy-weight group.

**Table 1:** Univariate analysis: Socio-demographic factors

| Factors | Categories | Low back pain (LBP) |  | Total (%) within categories | X-squared | p-value* |
| --- | --- | --- | --- | --- | --- | --- |
|  |  | No | Yes (Row %) |  |  |  |
| Age | ≤30 | 84 | 33 (28.2) | 117 (19.7%) | 11.452 | **0.003** |
|  | 31-40 | 227 | 123 (35.1) | 350 (59.1%) |  |  |
|  | 41-59 | 65 | 61 (48.4) | 126 (21.2%) |  |  |
| Gender | Female | 158 | 93 (37.1) | 251 (42.3%) | 0.012 | 0.911 |
|  | Male | 218 | 124 (36.3) | 342 (57.7) |  |  |
| BMI | Normal | 198 | 97 (32.9) | 295 (49.7%) | 7.707 | **0.021** |
|  | Obese | 17 | 21 (55.3) | 38 (6.5%) |  |  |
|  | Overweight | 161 | 99 (38.1) | 260 (43.8%) |  |  |
| Marital Status | Married | 304 | 193 (38.8) | 497 (83.8%) | 6.052 | **0.014** |
|  | Unmarried | 72 | 24 (25.0) | 96 (16.2%) |  |  |
| Crowding | ≤1.5 | 203 | 114 (36.0) | 317 (53.5%) | 0.133 | 0.936 |
|  | 1.5-2.0 | 118 | 71 (37.6) | 189 (31.9%) |  |  |
|  | 2+ | 55 | 32 (36.8) | 87 (14.6%) |  |  |
| Sleeping mattress | Firm bed | 316 | 186 (37.1) | 502 (84.7%) | 0.181 | 0.670 |
|  | Foam bed | 60 | 31 (34.7) | 91 (15.3%) |  |  |

\*p-value is calculated from chi-square test

**Table 2:** Univariate analysis: Behavioral and job related factors

| Factors | Categories | Low back pain (LBP) |  | Total (%) within categories | X-squared | p-value* |
| --- | --- | --- | --- | --- | --- | --- |
|  |  | No | Yes (Row %) |  |  |  |
| Smoking habit |  | No | Yes (Row %) |  | 0.109 | 0.740 |
|  | No | 310 | 182 (37.0) | 492 (83.0%) |  |  |
|  | Yes | 66 | 35 (34.7) | 101 (17.0%) |  |  |
| Chronic illness | No | 306 | 156 (33.8) | 462 (77.9%) | 6.664 | **0.009** |
|  | Yes | 70 | 61 (46.6) | 131 (22.1%) |  |  |
| Physical Activity | Sedentary | 110 | 78 (41.5) | 188 (31.7%) | 3.062 | 0.216 |
|  | Light | 231 | 123 (34.7) | 354 (59.7%) |  |  |
|  | Moderate-Vigorous | 35 | 16 (31.4) | 51 (8.6%) |  |  |
| Job experience (years) | ≤5 | 141 | 51 (26.6) | 192 (32.4%) | 16.095 | **<0.001** |
|  | 6-10 | 126 | 73 (36.7) | 199 (33.6%) |  |  |
|  | 10+ | 109 | 93 (46.0) | 202 (34.0%) |  |  |
| Working hour/day | 8-9 | 224 | 109 (32.7) | 333 (56.2%) | 4.507 | **0.033** |
|  | >9 | 152 | 108 (41.5) | 260 (43.8%) |  |  |

\*p-value is calculated from chi-square test

**Table 2:** Result from logistic regression model

| Factors | Reference | AOR (95%CI) | LCL | UCL | p-value |
| --- | --- | --- | --- | --- | --- |
| Age 31- 40 years | ≤30 years | 1.32 | 0.83 | 2.13 | 0.241 |
| Age 41-59 years | ≤30 years | **2.11** | **1.21** | **3.74** | **0.009** |
| BMI-cat- obese | Normal | **2.06** | **1.01** | **4.21** | **0.046** |
| BMI-cat-overweight | Normal | 1.14 | 0.79 | 1.63 | 0.484 |
| Office hours-extended (>9 hours) | Regular (8-9 hours) | **1.42** | **1.00** | **2.00** | **0.049** |
| Smoking habit- yes | No | 0.76 | 0.47 | 1.21 | 0.257 |
| Chronic illness- yes | No | 1.46 | 0.95 | 2.26 | 0.085 |
| Physical activity- light | Sedentary | 0.72 | 0.49 | 1.05 | 0.084 |
| Physical activity- moderate to vigorous | Sedentary | 0.057 | 0.28 | 1.12 | 0.110 |

### Variables importance in Random forest model

Here we interpret variables importance by applying a Random Forest model when predicting LBP. There is a consensus that random forests rarely suffer from “overfitting,” which plagues many other models [24]. We used mean decrease accuracy to assess random forests and to examine them on the probability of affecting LBP. The result of the random forest model is displayed in Figure 3, which shows a mean decrease in accuracy for the factors. The overall accuracy by the training data is 77.42%. It appears that the top 3 most important variables are: duration of employment, age, and long working hours. Hence, occupational factors play an important role on the complaints of LBP

**Figure 3:**
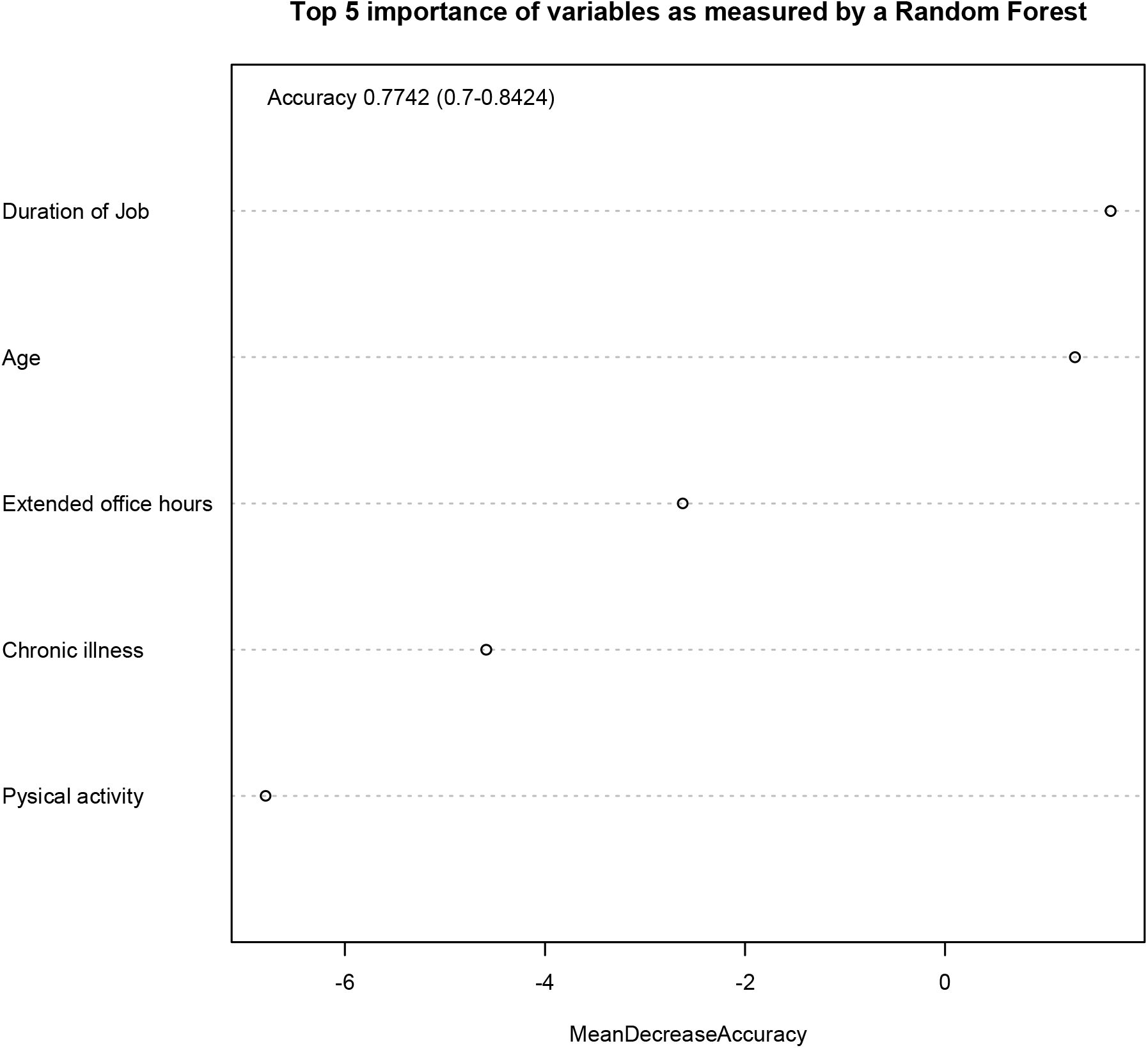
Random Forest Model.

## Discussion

The one-month prevalence of LBP among bank employees is 36.6%. Similar to our research, a cross-sectional study conducted in Kigali, Rwanda, found that 45.8% of bank staffs reported LBP during the one year [25]. Another study conducted in Nigeria found 12 months prevalence rate of office workers was 38.0% [26]. A cross-sectional study conducted in Pakistan among office workers found a 69.2% lifetime prevalence [27]. The one-year prevalence of LBP among information technology professionals in India was 51% [8]. In Malaysia, 37% of office workers experienced LBP in a year [28]. Among Thai university office workers, the three-months prevalence of LBP was 52.8% [29]. Additionally, the twelve-month prevalence of LBP in Australian adults and one-month period prevalence in the US general population was 67% and 39%, respectively [30-31]. Global point prevalence, one-month prevalence, and twelve-months prevalence of LBP among the adult population were estimated 12%, 23%, 38%, respectively [32]. Thus, the prevalence of LBP varied from 12% to 69%, and it depends on the type of questionnaire used, the period of prevalence, occupational provision, and lifestyle of the target population along with the location of the study.

There are many risk factors identified among the employees. Age is a common and substantial factor in developing LBP [33]. Our study found a higher number of older employees complained LBP than younger employees. Marital status also plays a role in developing LBP. A study conducted on the Iranian population also found similar results [34]. It may be because most married people belong to an older age group and so they complain about LBP.

This study’s results showed that regularly maintaining a long office hour is positively associated with increased complaints of LBP. A study conducted in Denmark also suggested that those employees who spend more time sitting down at the office caught LBP at a higher rate than those who spent less time in sitting positions at the office [35]. A similar finding was reported in another study conducted among university employees in Qatar [3]. The bank employees who were working for more than 10 years complained about LBP more than those who were working for less than 5 years. Similarly, an increased prevalence of LBP was found for office workers with many years of employment [17, 35-36].

We found that obesity has a strong association with the complaints of LBP. A meta-analysis also reported that obesity was associated with an increased prevalence of LBP in the past 12 months [37]. The gender difference was not evident in our study regarding the prevalence of complaining low back pain among office employees. But a meta-analysis suggests the prevalence of low back pain was stronger for women than for men [37]. Few studies suggest that regular physical activity can reduce LBP [21, 38]. In our study, the results show an association between physical activity and LBP at a 10% significance level, and the random forest method also identified the factor as an important variable.

There are many strengths to this study. We described gaps in the complaints of LBP from full-time bank employees. The study participants were homogenous because of their job nature, sitting arrangement, and the environment of the bank office was almost the same. Limitations include the self-reported data collected through study participant questionnaires. LBP differences observed in this paper cannot be interpreted as causative because of the consideration of the cross-sectional study design. In a future goal, a multi-professional study may help determine the overall generalizability of our results to the full-time employee as a whole.

## Conclusion

We found a high burden of low back pain among bank employees in Dhaka City. Studies revealed that long working hours and many years of employment are associated with LBP. The study showed a high prevalence in the 41 to 59-year-old age group, indicating that it is a common condition among older adults. Moreover, our findings add to the evidence on the importance of obesity about low back pain. Taking appropriate physical activity, taking a short break from the sitting position at the office, and doing body stretching at the office could be a low-cost solution for office workers.

## Data Availability

http://individual.utoronto.ca/ahmed_3/index_files/data/data.html

## Disclosure

### Ethical Consideration

We explained the aim and objectives of the research to the full-time bank employees. After having the verbal agreement from the participant, we distributed a written informed consent form and a questionnaire. The ethical committee of the Bangladesh University of Professionals (2019/273) and IRB of North South University (NSU-IRB-2019/54) approved the study.

### Consent to Publish

Not applicable.

### Availability of data

Click here for the data file http://individual.utoronto.ca/ahmed_3/index_files/data/data.html.

### Author’s contributions

MA and AH participated in study conception, design and coordination of the manuscript. GUA reviewed the manuscript and helped to draft the manuscript. AH also performed statistical analysis and helped to draft the manuscript. All authors approved the final manuscript.

## Acknowledgements

All the authors acknowledge the participants for providing us the information to conduct the study. We also thank Sheneeza Khan for assistance with English language usage, grammar, and spelling that greatly improved the manuscript.

## References

1. Vos T, Abajobir AA, Abate KH, Abbafati C, Abbas KM, Abd-Allah F, et al. Global, regional, and national incidence, prevalence, and years lived with disability for 328 diseases and injuries for 195 countries, 1990–2016: a systematic analysis for the Global Burden of Disease Study 2016. The Lancet. 2017 Sep 16; 90(10100):1211–59.

2. Hahne AJ, Ford JJ, Surkitt LD, Richards MC, Chan AYP, Thompson SL, et al. Specific treatment of problems of the spine (STOPS): Design of a randomized controlled trial comparing specific physiotherapy versus advice for people with subacute low back disorders. BMC Musculoskelet Disord. 2011;12(1):104.

3. Hanna F, Daas RN, El-Shareif TJ, Al-Marridi HH, Al-Rojoub ZM, Adegboye OA. The Relationship Between Sedentary Behavior, Back Pain, and Psychosocial Correlates Among University Employees. Front Public Heal. 2019 Apr 9; 7(APR):80.

4. Lotters F, Burdorf A. Prognostic Factors for Duration of Sickness Absence due to Musculoskeletal Disorders. Clin J Pain. 2006 Feb; 22(2):212–21.

5. Urhonen T, Lie A, Aamodt G. Associations between long commutes and subjective health complaints among railway workers in Norway. Prev Med Reports J. 2016; 4:490–495.

6. Andersen JH, Gaardboe O, Anderson SP, Oakman J, Article O, Arvidsson I, et al. National Institute for Working Life Ergonomic Expert Committee Document No 1Visual Display Unit Work and … Committee Document No 1 Visual Display Unit Work and Upper Extremity Musculoskeletal Disorders A Review of Epidemiological Findings. Appl Ergon. 2015;7(1):37–41.

7. Ayanniyi O, Ukpai B, Adeniyi A. Differences in prevalence of self-reported musculoskeletal symptoms among computer and non-computer users in a Nigerian population: a cross-sectional study. BMC Musculoskelet Disord. 2010 Dec 6 [cited 2019 Sep 5];11(1):177.

8. Hameed S. Prevalence of Work-Related Low Back Pain Among The Information Technology Professionals In India-A Cross-Sectional Study. Int J Sci Technol Res. 2013;2(7).

9. Campos-Fumero A, Delclos GL, Douphrate DI, Felknor SA, Vargas-Prada S, Serra C, et al. Low back pain among office workers in three Spanish-speaking countries: findings from the CUPID study. Inj Prev. 2017 Jun;23(3):158–64.

10. Ye S, Jing Q, Wei C, Lu J. Risk factors of non-specific neck pain and low back pain in computer-using office workers in China: a cross-sectional study. BMJ Open. 2017 Apr 11;7(4):e014914.

11. Janwantanakul P, Pensri P, Jiamjarasrangsri V, Sinsongsook T. Prevalence of self-reported musculoskeletal symptoms among office workers. Occup Med (Chic Ill). 2008;58(6):436–8.

12. Janwantanakul P, Sihawong R, Sitthipornvorakul E, Paksaichol A. A screening tool for non-specific low back pain with disability in office workers: A 1-year prospective cohort study Rehabilitation, physical therapy, and occupational health. BMC Musculoskelet Disord. 2015;16(1):1–8.

13. P. J, P. P, P. M, W. J. Development of a risk score for low back pain in office workers -a cross-sectional study. BMC Musculoskelet Disord. 2011;12.

14. Sihawong R, Janwantanakul P, Jiamjarasrangsi W. A prospective, cluster-randomized controlled trial of exercise program to prevent low back pain in office workers. Eur Spine J [Internet]. 2014 Apr 4 [cited 2019 Sep 6];23(4):786–93.

15. Waongenngarm P, Rajaratnam BS, Janwantanakul P. Perceived body discomfort and trunk muscle activity in three prolonged sitting postures. J Phys Ther Sci [Internet]. 2015;27(7):2183–7.

16. Dunstan DW, Wiesner G, Eakin EG, Neuhaus M, Owen N, LaMontagne AD, et al. Reducing office workers’ sitting time: rationale and study design for the Stand Up Victoria cluster randomized trial. BMC Public Health [Internet]. 2013 Dec 9;13(1):1057.

17. Leitaru N, Kremers S, Hagberg J, Björklund C, Kwak L. Associations Between Job-Strain, Physical Activity, Health Status, and Sleep Quality Among Swedish Municipality Workers. J Occup Environ Med [Internet]. 2019 Feb;61(2):e56–60.

18. Fatoye F, Gebrye T, Odeyemi I. Real-world incidence and prevalence of low back pain using routinely collected data [Internet]. Vol. 39, Rheumatology International. Springer Berlin Heidelberg; 2019 [cited 2019 Sep 5]. p. 619–26.

19. Hossain A. Prevalence and Occupational Factors Associated with Low Back Pain Among the Female Garment Workers: A Cross-Sectional Study in Bangladesh. Glob Perspect Med Sci. 2018;2(1):8.

20. Sanjoy SS, Ahsan GU, Nabi H, Joy ZF, Hossain A. Occupational factors and low back pain: a cross-sectional study of Bangladeshi female nurses. BMC Res Notes. 2017;10(1):1–6.

21. Hossain MD, Aftab A, Al Imam MH, Mahmud I, Chowdhury IA, Kabir RI, et al. Prevalence of work-related musculoskeletal disorders (WMSDs) and ergonomic risk assessment among readymade garment workers of Bangladesh: A cross-sectional study. Guo NL, editor. PLoS One [Internet]. 2018 Jul 6 [cited 2019 Aug 28];13(7):e0200122.

22. Eriksen HR, Ihleb×k C, Ursin H. A scoring system for subjective health complaints (SHC). J Public Heal. 1999;1:63–72.

23. Mendes M de A, da Silva I, Ramires V, Reichert F, Martins R, Ferreira R, et al. Metabolic equivalent of task (METs) thresholds as an indicator of physical activity intensity. Zagatto AM, editor. PLoS One. 2018 Jul 19 [cited 2019 Aug 26];13(7):e0200701.

24. Robin Genuer, Jean-Michel Poggi, Christine Tuleau-Malot. Variable selection using Random Forests. Pattern Recognition Letters, 2010, 31 (14), pp.2225–2236. hal-00755489.

25. Kanyenyeri L, Asiimwe B, Mochama M, Nyiligira J, Habtu M. Prevalence of Back Pain and Associated Factors among Bank Staff in Selected Banks in Kigali, Rwanda: A Cross-Sectional Study. Heal Sci J. 2017;11(3):1–7.

26. Omokhodion FO, Sanya AO. Risk factors for low back pain among office workers in Ibadan, Southwest Nigeria. Occup Med (Chic Ill). 2003;53(4):287–9.

27. Arslan SA, Hadian MR, Olyaei G, Bagheri H, Yekaninejad MS, Ijaz S, et al. Prevalence and Risk Factors of Low Back Pain Among the Office Workers of King Edward Medical University Lahore, Pakistan. Phys Treat - Specif Phys Ther. 2017;6(3):161–8.

28. Damanhuri Z, Zulkifli A, Lau ACT, Zainuddin H. Low Back Pain Among Office Workers in a Public University in Malaysia. Int J Public Heal Clin Sci. 2014;1(1).

29. Chaiklieng S, Suggaravetsiri P, Stewart J. Incidence rate and risk factors associated with low back pain among university office workers in Thailand. 2015;(August):1–9.

30. Ferguson SA, Merryweather A, Thiese MS, Hegmann KT, Lu M-L, Kapellusch JM, et al. Prevalence of low back pain, seeking medical care, and lost time due to low back pain among manual material handling workers in the United States. BMC Musculoskelet Disord. 2019 Dec 22;20(1):243.

31. Papageorgiou AC, Croft PR, Ferry S, Jayson MI, Silman AJ. Estimating the prevalence of low back pain in the general population. Evidence from the South Manchester Back Pain Survey. Spine (Phila Pa 1976). 1995 Sep 1; 20(17):1889–94.

32. Manchikanti L, Singh V, Falco FJE, Benyamin RM, Hirsch JA. Epidemiology of Low Back Pain in Adults. Neuromodulation Technol Neural Interface. 2014; 17(S2):3–10.

33. Wong AY, Karppinen J, Samartzis D. Low back pain in older adults: risk factors, management options, and future directions. Scoliosis spinal Disord. 2017; 12:14.

34. Biglarian A, Seifi B, Bakhshi E, Mohammad K, Rahgozar M, Karimlou M, et al. Low back pain prevalence and associated factors in Iranian population: Findings from the national health survey, Pain Research and Treatment. 2012.p. 1–5.

35. Gupta N, Christiansen CS, Hallman DM, Korshøj M, Carneiro IG, Holtermann A. Is Objectively Measured Sitting Time Associated with Low Back Pain? A Cross-Sectional Investigation in the NOMAD study.Dorner TE, editor. PLoS One. 2015 Mar 25;10(3):e0121159.

36. Lis AM, Black KM, Korn H, Nordin M. Association between sitting and occupational LBP. Eur Spine J. 2007 Feb;16(2):283–98.

37. Rahman Shiri, Jaro Karppinen, Päivi Leino-Arjas, Svetlana Solovieva, Eira Viikari-Juntura The Association Between Obesity and Low Back Pain: A Meta-Analysis, American Journal of Epidemiology, December 11, 2009, Vol. 171, No. 2 DOI: 10.1093/aje/kwp356.

38. Gordon R, Bloxham S. A Systematic Review of the Effects of Exercise and Physical Activity on Non-Specific Chronic Low Back Pain. Healthcare. 2016 Apr 25;4(2):22.

